# Blood-based transcriptomic classification of lung cancer: a leakage-free nested cross-validation framework with LASSO

**DOI:** 10.64898/2026.07.11.26357823

**Authors:** Sumeyye Bakim, Nurten Urlu Özalan, Elif Gülbahçe Mutlu, Vahdettin Demir, Erdi Gülbahçe

## Abstract

Peripheral whole-blood gene expression profiling offers a minimally invasive route to lung cancer detection, but high-dimensional transcriptomic data are prone to optimistic bias when preprocessing and model selection are not properly separated from performance evaluation. We applied *ℓ*_1_-penalised (LASSO) logistic regression to 303 peripheral whole-blood microarray profiles (123 lung cancer cases and 180 healthy controls; Gene Expression Omnibus accession GSE252168; Illumina HumanHT-12 v4) within a leakage-free nested cross-validation framework (5 outer and 3 inner folds), in which all data-dependent steps—imputation, univariate feature screening by ANOVA *F*-test (*k* = 500), and standardisation—were confined strictly to training partitions. Statistical significance was assessed by permutation testing (*B* = 100), and feature selection stability was quantified across outer folds. LASSO was compared with ridge logistic regression, linear support vector machines, and random forest under the same framework. The LASSO model identified a sparse 29-probe signature with a pooled out-of-fold area under the ROC curve (AUC) of 0.990 (nested estimate 0.989 ± 0.015), accuracy 97.4%, sensitivity 94.3%, and specificity 99.4% at a 0.50 threshold; permutation testing confirmed significance (*p* = 0.0099). Six probes, including *CDC42, U2AF1*, and *RPS15A*, were selected in all five outer folds, forming a stable core, and all classifiers exceeded AUC 0.987, indicating a strong, algorithm-independent signal. A leakage-free nested cross-validation framework enables unbiased performance estimation and reproducible feature selection in blood-based lung cancer classification. The 29-probe panel is an internally validated candidate requiring prospective, multicentre external validation before clinical use.

## 1. Introduction

Lung cancer is the most commonly diagnosed cancer and the leading cause of cancerrelated mortality worldwide, accounting for approximately 2.5 million new cases and 1.8 million deaths in 2022 (Bray et al., 2024; Sung et al., 2021). Despite advances in systemic therapy, the five-year survival rate remains below 25% in most populations, principally because the majority of patients are diagnosed at advanced stages when curative intervention is no longer feasible (Siegel et al., 2024). Low-dose computed tomography (LDCT) screening has demonstrated a mortality reduction of up to 20% among high-risk individuals (National Lung Screening Trial Research Team, 2011), yet its widespread implementation is constrained by cost, limited accessibility, radiation exposure, and high false-positive rates that lead to unnecessary invasive procedures. These limitations have motivated a growing body of research into minimally invasive molecular biomarkers that could complement or refine existing screening strategies.

Blood-based transcriptomic profiling has emerged as a promising approach to cancer detection, reflecting the fact that systemic interactions between a tumour and the host immune system produce measurable changes in peripheral blood gene expression (Liew et al., 2006; Twine et al., 2003). Several studies have reported mRNA signatures in whole blood capable of discriminating lung cancer patients from healthy controls. Rotunno et al. (2011) identified an eight-gene signature from peripheral whole-blood samples with an AUC of 0.81 for stage I lung adenocarcinoma, while Showe et al. (2009) reported a 29-gene classifier achieving 86% accuracy in an independent validation cohort. More recently, multi-cohort meta-analyses have leveraged large-scale blood gene expression data to derive diagnostic scores validated across heterogeneous populations, collectively demonstrating that peripheral blood carries an informative transcriptomic signal for lung cancer; however, none of the proposed signatures has yet been translated into clinical practice (Zheng et al., 2025).

A recurring methodological challenge in this literature is the high-dimensional nature of gene expression data, where the number of measured features (*p*) vastly exceeds the number of available samples (*n*). Under such conditions, standard classification approaches are prone to overfitting, and reported performance metrics may be inflated by subtle forms of information leakage during model selection and evaluation (Ambroise and McLachlan, 2002; Simon et al., 2003). Regularised methods, in particular the least absolute shrinkage and selection operator (LASSO) (Tibshirani, 1996), address both dimensionality and interpretability by simultaneously performing coefficient estimation and variable selection. The LASSO has become one of the most widely adopted tools for constructing parsimonious gene expression classifiers (Hastie et al., 2009), yet its application in high-dimensional settings raises important methodological questions: how should the regularisation parameter be tuned without introducing selection bias? How sensitive is the selected feature set to the data partition? And how does LASSO compare, in both discrimination and sparsity, to alternative regularisation strategies? More broadly, comprehensive simulation studies have demonstrated that the choice of classifier, transformation strategy, and dataset characteristics critically influence classification accuracy in gene expression settings (Zararsiz et al., 2017), underscoring the need for systematic multi-method comparison within rigorous validation frameworks.

These questions have direct consequences for model evaluation and selection in practice. Varma and Simon (2006) demonstrated that using the same cross-validation loop for both hyperparameter selection and performance estimation introduces an optimistic bias whose magnitude depends on the ratio *n/p* and the strength of the classification signal. Cawley and Talbot (2010) further showed that this bias leads to the selection of overly complex models. The recommended remedy is nested (double) cross-validation, which employs separate inner and outer resampling loops for hyperparameter tuning and performance evaluation, respectively. Despite this well-established guidance, many gene expression classification studies continue to rely on single-loop procedures or leave-one-out cross-validation without correcting for the resulting bias, making it difficult to assess the true generalisability of proposed classifiers.

In this study, we develop a blood-based gene expression classifier for lung cancer detection within a methodologically rigorous framework. We apply *ℓ*_1_-penalised logistic regression to publicly available whole-blood microarray data from 303 peripheral blood samples (GSE252168) within a leakage-free nested cross-validation pipeline, in which all data-dependent preprocessing steps (imputation, univariate feature screening, and standardisation) are performed exclusively on the training partition at each level of the nested procedure, thereby eliminating information leakage as a source of optimistic bias (Ambroise and McLachlan, 2002). We complement the primary analysis with a systematic comparison of four classification methods (LASSO, ridge logistic regression, linear SVM, and random forest), a permutation test for statistical significance, a comprehensive assessment of feature selection stability, and an analysis of the pairwise correlation structure of retained probes. The principal objectives of this work are twofold: to identify a compact, blood-based gene expression signature capable of discriminating lung cancer cases from healthy controls; and to provide a reproducible methodological template for rigorous penalised classification in high-dimensional genomic settings. A companion study of the same cohort investigates the derivation of a qRT-PCR-compatible panel and the cohort–batch confound in detail.

## 2. Materials AND Methods

### 2.1. Data source and preprocessing

The analysis used publicly available gene expression data from the Gene Expression Omnibus (GEO) repository under accession number GSE252168 (Edgar et al., 2002; Skogholt et al., 2024). The dataset comprises *n* = 303 peripheral whole-blood samples: 123 lung cancer patients recruited at St Olavs University Hospital, Trondheim, Norway, and 180 healthy controls drawn from two population-based biobanks: 118 participants from the HUNT study (Tempus tubes) and 62 samples from 34 women in the Norwegian Women and Cancer cohort (NOWAC; PAXgene tubes with technical replicates). Whole-blood gene expression was profiled using the Illumina HumanHT-12 v4 Expression BeadChip platform (GPL10558), measuring over 47,000 transcripts, of which *p*_0_ = 30,715 probes passed quality filtering and were retained in the normalised expression matrix. The original investigators performed background correction, quantile normalisation, and probe-level batch correction to account for systematic expression differences between the PAXgene and Tempus collection systems (Skogholt et al., 2024). Sample-level annotations were available in the public repository but were not used in the present analysis, which focuses exclusively on gene expression-based classification.

All data-dependent preprocessing steps were embedded within a single scikit-learn Pipeline object (Pedregosa et al., 2011) and fitted exclusively on the training data within each cross-validation fold, following established recommendations for avoiding information leakage in high-dimensional classification (Ambroise and McLachlan, 2002; Simon et al., 2003). The pipeline proceeded in three sequential stages. First, probe-level missing values were imputed using the training-fold mean. Second, a univariate ANOVA *F*-test filter retained the top *k* = 500 probes with the strongest marginal association with the binary outcome, equivalent to a two-sample *t*-test in the binary case. The threshold *k* = 500 was chosen to be deliberately conservative relative to the theoretical lower bound of *k* = *O*(*n/* log *n*) ≈ 53 suggested by the sure independence screening literature (Fan and Lv, 2008) for *n* = 303, ensuring that probes with moderate marginal effects were not prematurely excluded while still achieving a tenfold reduction in dimensionality (from 30,715 to 500) prior to penalised regression. This choice is consistent with common practice in gene expression classification, where pre-screening thresholds of several hundred to a few thousand features are widely adopted (Haury et al., 2011). Third, retained features were standardised to zero mean and unit variance, with test-fold features transformed using training-fold parameters. All preprocessing steps were fitted exclusively on the training partitions at each resampling step to prevent any potential information leakage.

### 2.2. Classification methods

The primary classifier was *ℓ*_1_-penalised (LASSO) logistic regression (Tibshirani, 1996). Under the logistic model, the class-membership probability is

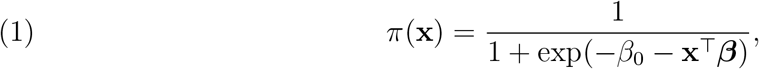

and the LASSO estimator maximises the penalised log-likelihood

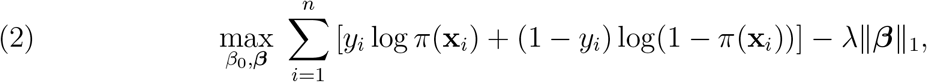

where 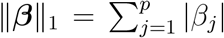 and *λ* ≥0 controls regularisation strength. For computational convenience, regularisation was parametrised as *C* = 1*/λ*, with the candidate grid *C* ∈ {0.01, 0.05, 0.10, 0.50, 1.00}. The L1 penalty induces sparsity by shrinking coefficients to exactly zero, thereby performing simultaneous estimation and feature selection. The solution path was computed using the SAGA solver (Defazio et al., 2014) with a maximum of 5000 iterations; convergence was reached well before this limit in all cases.

Three alternative classifiers were included for systematic comparison. Ridge logistic regression (Hoerl and Kennard, 1970) replaces the L1 penalty with 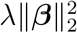, shrinking all coefficients toward zero without inducing exact sparsity; it was tuned over *C* ∈ {0.1, 1.0, 10.0}. A linear support vector machine (SVM) (Cortes and Vapnik, 1995) seeks the maximum-margin separating hyperplane with soft-margin parameter *C* ∈ {0.1, 1.0}. For both methods, *C* was selected via the inner cross-validation loop. Random forest (Breiman, 2001) was included as a nonlinear comparator; it constructs an ensemble of 200 decision trees (maximum depth 10) trained on bootstrap samples with random feature subsets. Random forest hyperparameters were fixed a priori and were not optimised via inner cross-validation, as the method was included primarily as a nonlinear baseline; only outer-fold performance was evaluated.

### 2.3. Validation and statistical inference

Unbiased estimation of generalisation performance requires separating hyperparameter selection from model evaluation. We implemented nested (double) cross-validation (Varma and Simon, 2006; Cawley and Talbot, 2010) consisting of an outer stratified 5-fold loop for performance assessment and an inner stratified 3-fold loop for hyperparameter tuning. For each outer fold *k*, the inner loop evaluated all candidate hyperparameter values on the training partition and selected the configuration maximising mean ROC-AUC. The model was then retrained on the full outer training set with the selected hyperparameters and evaluated on the held-out test fold. Critically, the entire preprocessing pipeline was refitted independently within each training partition at both the inner and outer levels, ensuring that no information from test observations influenced any preprocessing step. Algorithm 1 summarises this procedure.

In addition to fold-level AUC values, a pooled out-of-fold (OOF) AUC was computed by aggregating the predicted probabilities from all five outer folds into a single vector and evaluating it against the true labels, providing a single global performance estimate in which each observation contributes only to its held-out prediction. Binary classification metrics, including accuracy, sensitivity, specificity, F1 score, and the Matthews correlation coefficient (MCC) (Matthews, 1975), were computed from OOF predictions at a decision threshold of 0.50.

#### Algorithm 1

Leakage-free nested cross-validation

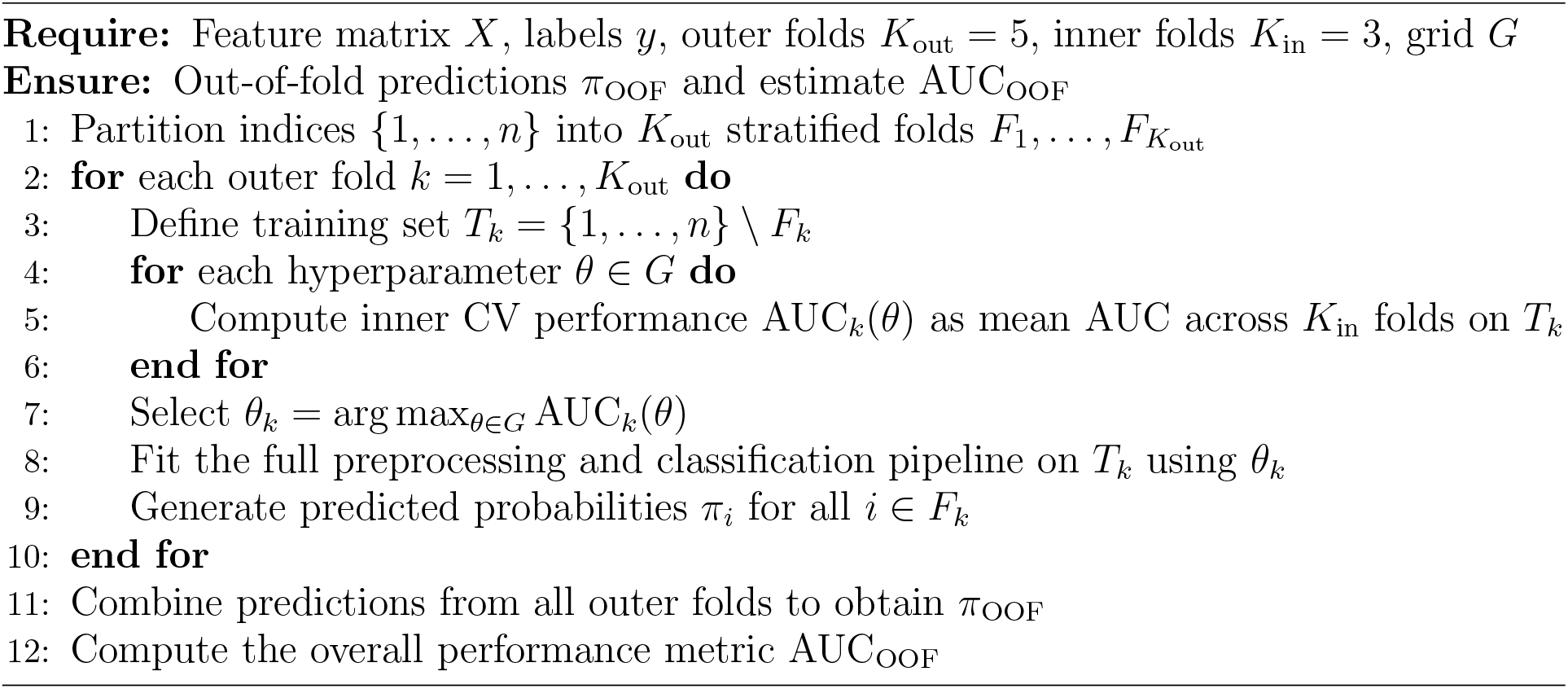

Statistical significance was assessed via a permutation test (Good, 2005; Ojala and Garriga, 2010). Class labels were randomly shuffled *B* = 100 times while keeping the feature matrix intact, and the full nested cross-validation pipeline was refit under each permutation to obtain a null distribution of AUC values. The *p*-value was computed as

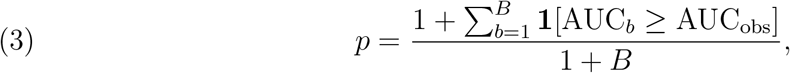

where AUC_*b*_ is the AUC under permutation *b*, AUC_obs_ is the observed AUC, and **1**(·) is the indicator function. Following the convention of Ojala and Garriga (2010), one is added to both numerator and denominator to avoid a zero *p*-value when the observed statistic exceeds all permuted values. The test used the modal regularisation parameter (*C* = 0.10) held fixed across permutations. Although 100 permutations provide limited resolution, the observed AUC exceeded all permuted values, yielding the smallest attainable value under this design (*p* = 1*/*(1 + *B*) = 0.0099).

### 2.4. Feature stability assessment

Feature stability was quantified by tracking which probes received non-zero LASSO coefficients across the five outer folds. For each probe *j*, the selection frequency was defined as

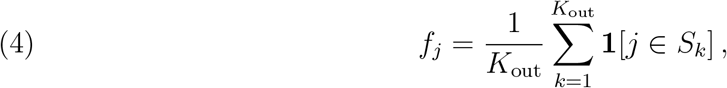

where *K*_out_ is the number of outer folds and *S*_*k*_ is the set of features selected in fold *k*. Pairwise overlap between folds was summarised using the Jaccard similarity index

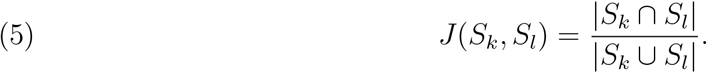

The mean Jaccard index across all pairwise fold comparisons was used as a global measure of feature selection stability (Meinshausen and Bühlmann, 2010). A final deployable model was constructed by fitting the LASSO pipeline on the entire dataset using the modal regularisation parameter (Varma and Simon, 2006), and feature stability was additionally characterised by examining the pairwise Pearson correlation structure of the retained probes to contextualise the known tendency of *ℓ*_1_-penalised estimators to select one representative from groups of correlated predictors (Zou and Hastie, 2005). All analyses were conducted in Python 3.10 using scikit-learn (Pedregosa et al., 2011), and all random seeds were fixed for reproducibility.

## 3. Results

The LASSO-penalised logistic regression model, trained and evaluated within a leakage-free nested cross-validation framework on 303 peripheral whole-blood transcriptomic profiles (123 lung cancer cases, 180 healthy controls), identified a sparse 29-probe signature with strong discriminative performance.

### 3.1. Nested cross-validation performance

Five-fold nested cross-validation yielded a mean outer-fold AUC of 0.989 (SD = 0.015), with individual fold values ranging from 0.960 to 1.000, indicating consistently strong discrimination regardless of the specific training–test partition (Table 1). The inner loop selected the regularisation parameter *C* independently within each outer fold; the modal value was *C* = 0.10, chosen in three of five folds, with the remaining folds selecting *C* = 0.50 and *C* = 0.05. The moderate variation in optimal *C* reflects expected hyperparameter uncertainty in high-dimensional settings, yet the uniformly high outer-fold AUC values confirm that performance was robust to this variation. Aggregating the OOF predicted probabilities across all five outer folds, the pooled OOF AUC was 0.990. A permutation test based on 100 random label shuffles produced a null AUC distribution centred at approximately 0.50; the observed AUC exceeded all permuted values (*p* = 0.0099), confirming that the selected probe features carry authentic predictive information about case–control status.

**TABLE 1.** Nested five-fold cross-validation results for the LASSO classifier.

| Fold | Optimal $C$ | Inner AUC | Outer AUC | $N$ selected |
| --- | --- | --- | --- | --- |
| 1 | 0.50 | 0.960 | 0.960 | 39 |
| 2 | 0.10 | 1.000 | 1.000 | 30 |
| 3 | 0.10 | 0.986 | 0.986 | 29 |
| 4 | 0.05 | 1.000 | 1.000 | 22 |
| 5 | 0.10 | 0.999 | 0.999 | 35 |
| Mean | — | 0.989 | 0.989 | 31.0 |

### 3.2. Classification performance at the decision threshold

At the decision threshold of 0.50, the pooled OOF predictions yielded an overall accuracy of 97.4%. The confusion matrix (Figure 1) shows that 179 of 180 control samples were correctly classified (specificity = 99.4%) and 116 of 123 cancer cases were correctly identified (sensitivity = 94.3%). Only seven cancer samples were misclassified as controls (false negatives), and a single control sample was incorrectly assigned as a case (false positive). The F1 score was 0.967 and the MCC was 0.946, both indicating strong agreement between predicted and true labels across both classes. The distribution of OOF predicted probabilities (Figure 2) shows control samples concentrated near zero and case samples predominantly exceeding 0.80, with minimal overlap in the intermediate region (0.40–0.60), demonstrating well-separated probability estimates.

**FIGURE 1.**
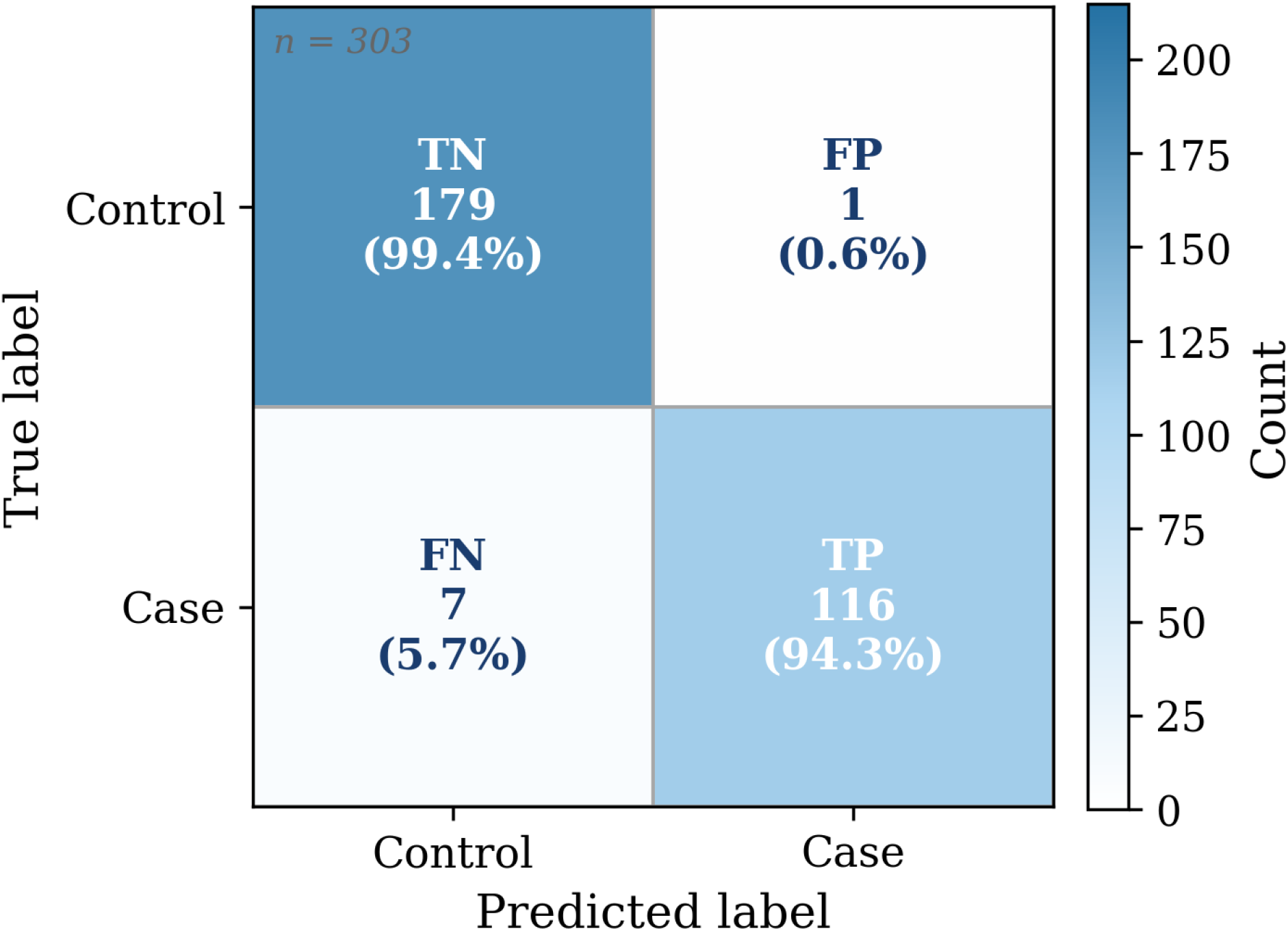
Confusion matrix of pooled out-of-fold predictions at decision threshold 0.50. Cell values show absolute counts and within-row percentages. TN, true negative; FP, false positive; FN, false negative; TP, true positive.

**FIGURE 2.**
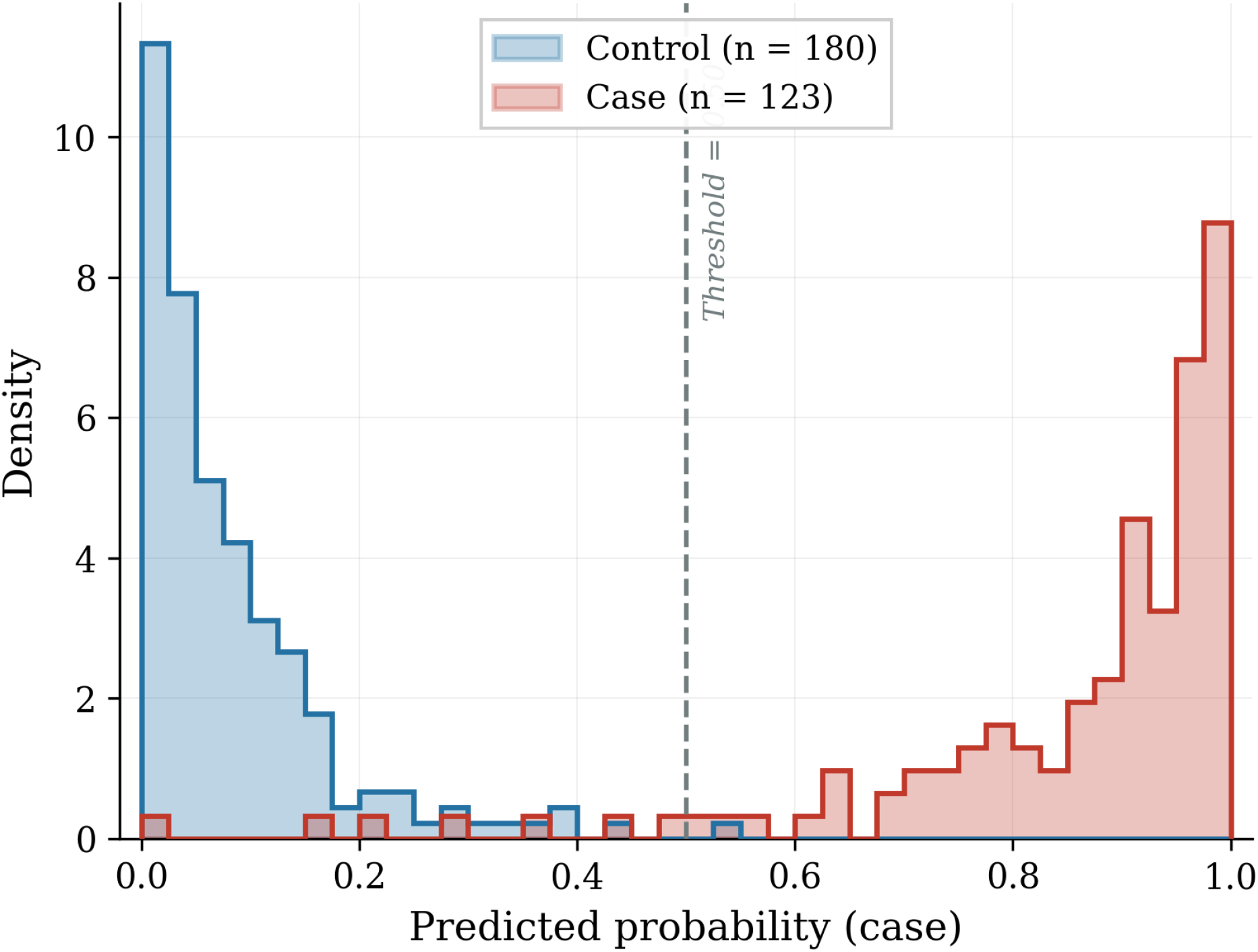
Distribution of out-of-fold predicted probabilities for control and case samples. The dashed vertical line indicates the decision threshold at 0.50.

### 3.3. Selected gene panel and feature stability

The final LASSO model, trained on the full dataset with the modal regularisation parameter (*C* = 0.10), retained 29 probes with non-zero coefficients. Of these, 20 could be mapped to annotated genes, while the remaining nine correspond to unannotated transcripts or predicted loci. The coefficient profile of the top 20 features, ranked by absolute magnitude, is displayed in Figure 3. A pronounced directional asymmetry was observed: 22 of 29 selected probes carried negative coefficients (relative downregulation in cancer), while seven had positive coefficients. Among annotated genes, *ENSA* (*β* = +0.574) exhibited the largest positive coefficient, followed by *TTN* (*β* = +0.458) and *CDC42* (*β* = +0.349). On the negative side, an unannotated probe (ILMN 1898691; *β* = −0.445) and *LOC100130715* (*β* = −0.315) displayed the strongest suppressive contributions, followed by *RPS15A* (*β* = −0.288). The top-ranked subset is presented in Table 2.

**TABLE 2.**
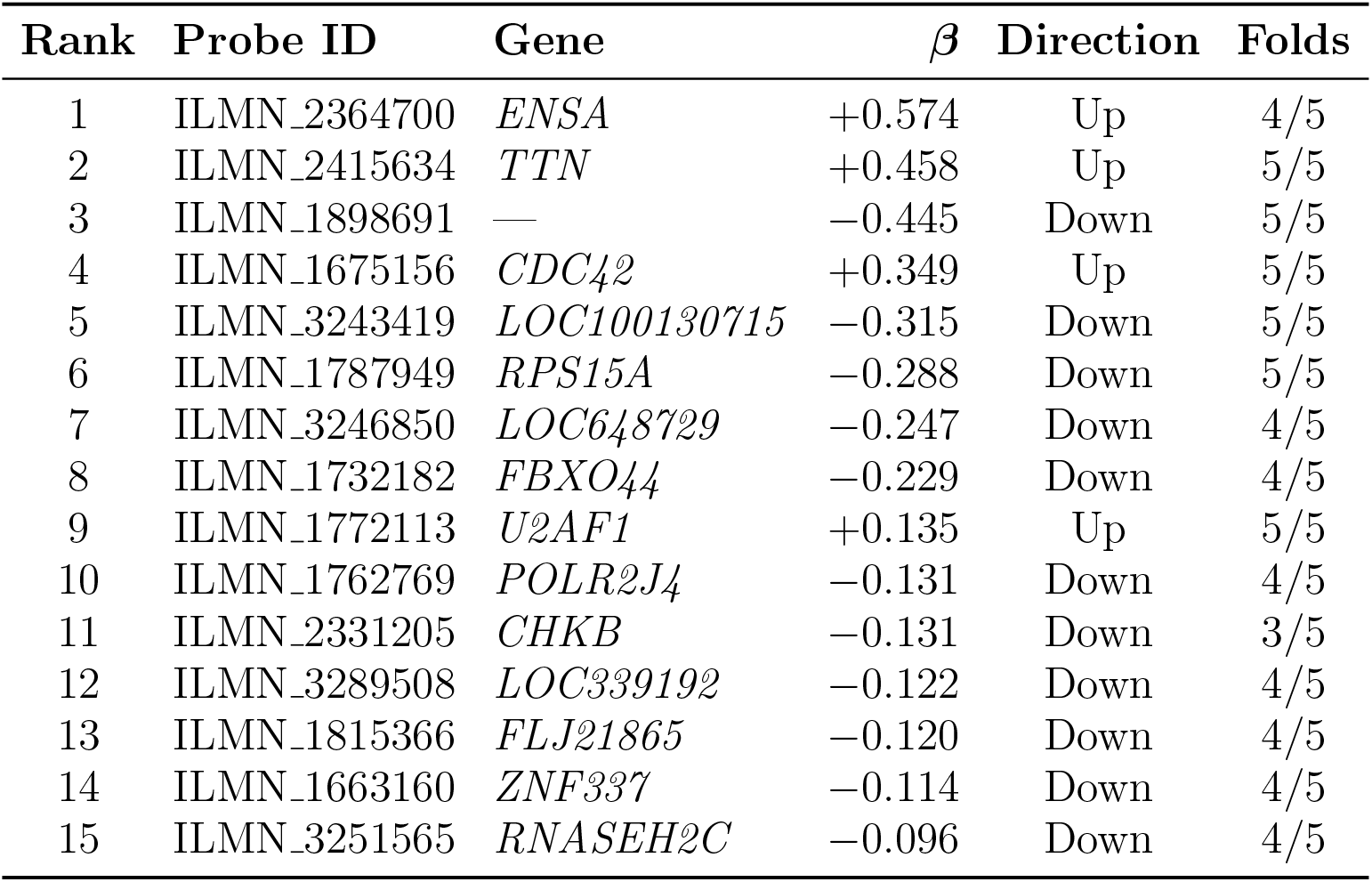
Top 15 features of the final LASSO model, ranked by |*β*|.

**FIGURE 3.**
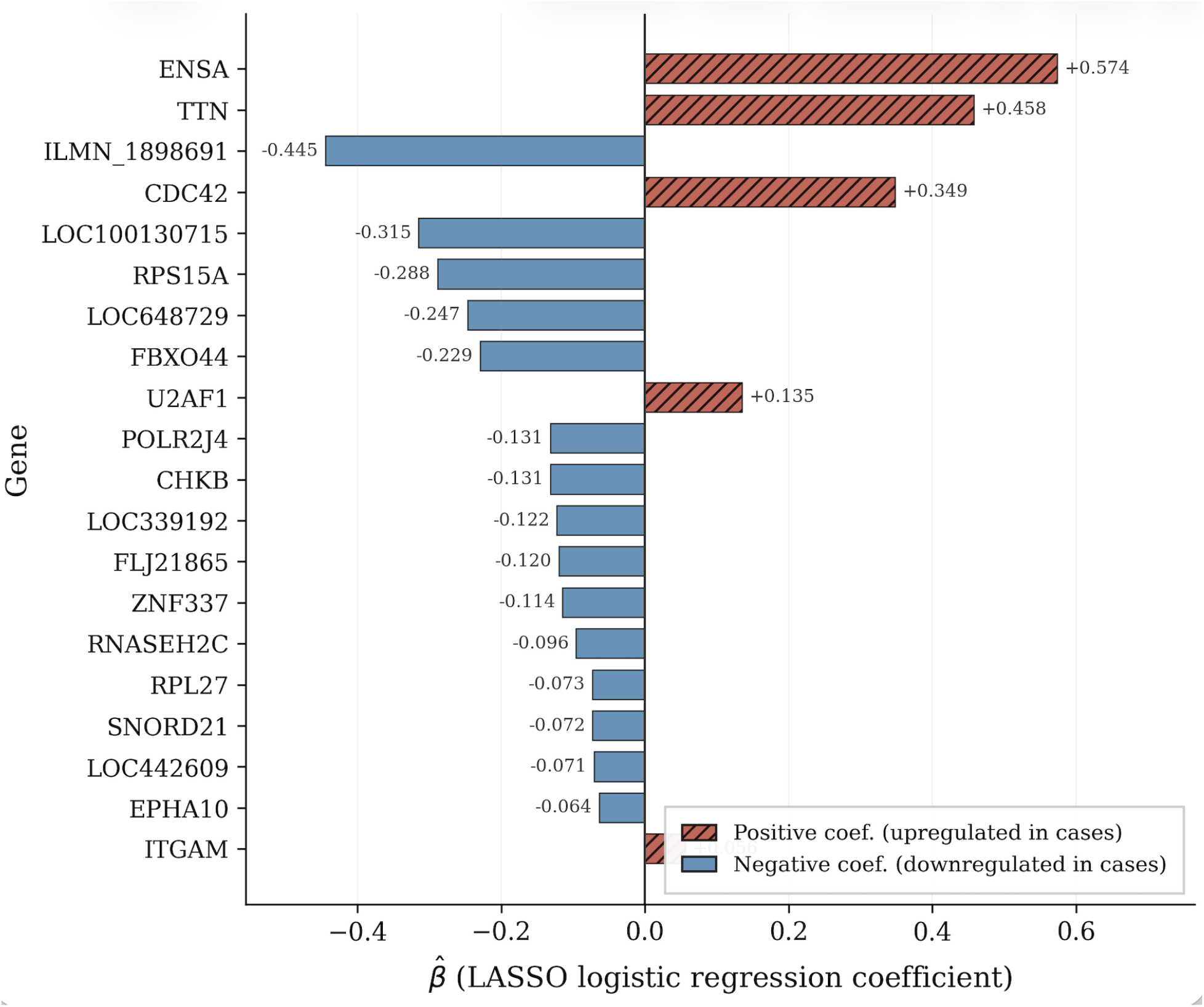
LASSO coefficients for the top 20 features ranked by 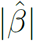. Positive coefficients indicate upregulation in cancer; negative coefficients indicate downregulation.

Feature stability was assessed by tracking probe selection across the five outer folds (Figure 4). Six probes formed a highly stable core, appearing in all five folds (selection frequency = 1.00): *CDC42, U2AF1, RPS15A*, ILMN 1898691, *TTN*, and *LOC100130715*.

**FIGURE 4.**
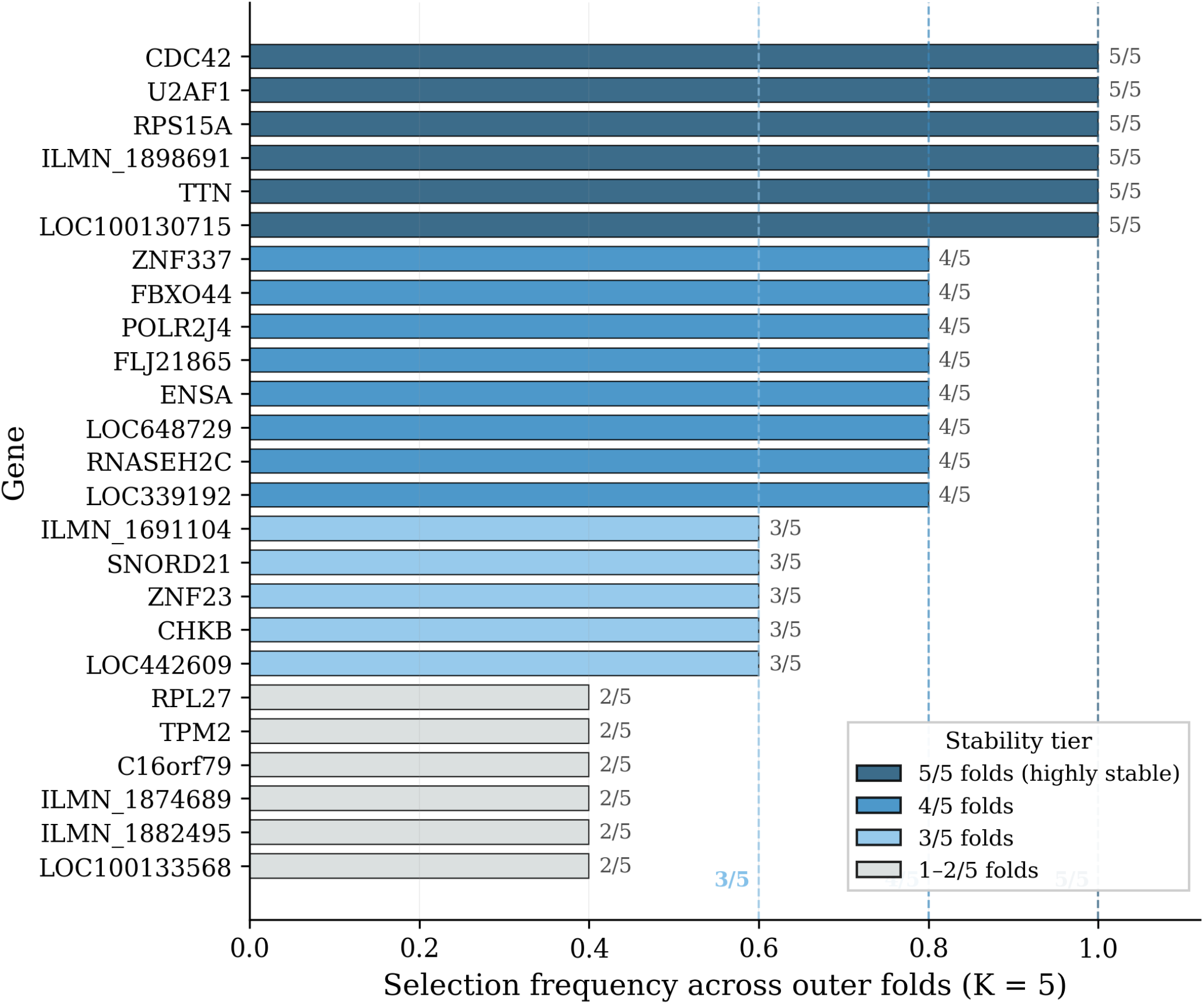
Feature selection frequency across the five outer cross-validation folds (top 25 probes). Colour indicates stability tier.

These span functionally diverse categories, including cell signalling (*CDC42*), mRNA splicing (*U2AF1*), translational machinery (*RPS15A*), and structural proteins (*TTN*), suggesting a multi-dimensional transcriptomic shift rather than a single pathway. An additional eight probes appeared in four of five folds, extending the reproducible core to 14 features. The mean pairwise Jaccard index across all fold comparisons was 0.401 (SD = 0.093), reflecting the expected variability of *ℓ*_1_-penalised solutions under correlated predictors. In total, 50 unique probes were selected at least once, of which 14 (28%) appeared in four or more folds.

### 3.4. Correlation structure of the selected gene panel

To characterise co-expression among the 29 retained probes, we computed the pairwise Pearson correlation matrix and applied hierarchical clustering (average linkage, correlation distance) (Figure 5). Two dominant co-expression modules were identified. The first, comprising most downregulated probes (including *CHKB, ZNF337, RNASEH2C, RPS15A, LOC648729, FBXO44, LOC100130715*, and ILMN 1898691), formed a block of moderate positive internal correlations. The second module grouped the upregulated probes *TTN, U2AF1, ITGAM, ENSA*, and *CDC42*, with a pronounced negative inter-cluster correlation that mirrors the asymmetric directionality of the LASSO coefficients. The mean absolute pairwise correlation was moderate (mean |*ρ*| = 0.498, median 0.480), with 172 of 406 probe pairs (42.4%) exceeding |*ρ*| = 0.50. Notably, two probes mapping to *RPS15A* and two mapping to *CHKB* were jointly retained and exhibited high within-pair correlations, illustrating the tendency of *ℓ*_1_-penalised estimators to retain multiple representatives from a correlated pair depending on the resampling split. In contrast, *CDC42*, a member of the stable core, showed comparatively low pairwise correlations, suggesting a robust, non-redundant signal.

**FIGURE 5.**
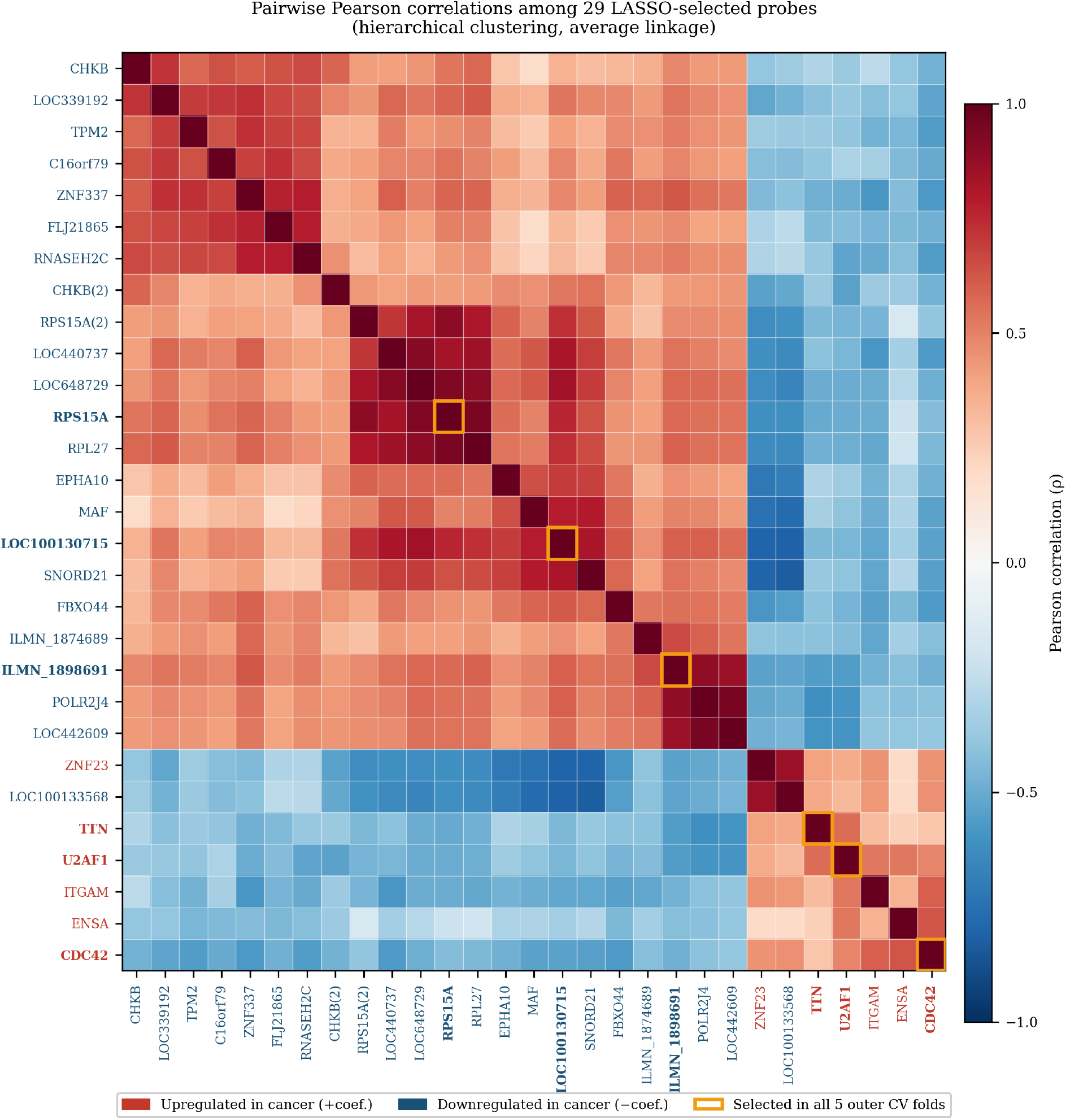
Pairwise Pearson correlation matrix of the 29 retained probes, ordered by hierarchical clustering. Bold labels indicate the six probes selected in all five outer folds. Mean |*ρ*| = 0.498; 172 of 406 pairs (42.4%) exceeded |*ρ*| = 0.50.

### 3.5. Comparison with alternative classification methods

The same leakage-free framework was applied to three alternative classifiers (Table 3, Figure 6). All four classifiers achieved uniformly high performance. Ridge regression yielded the highest mean AUC (0.999 ±0.002), followed by SVM (linear) (0.998 ±0.002), LASSO (0.989± 0.015), and random forest (0.987 ± 0.014). The differences across methods were small relative to within-method variability, indicating that the underlying signal is robust and not an artefact of any particular approach. However, ridge and SVM retained all input features with non-zero coefficients, producing dense models, whereas LASSO reduced the feature space to 29 probes, yielding a substantially more interpretable model amenable to biological annotation. The increased variability of the LASSO AUC (SD = 0.015 vs. 0.002 for ridge) is consistent with the known sensitivity of *ℓ*_1_-penalised estimators to correlated predictors. Random forest achieved comparable performance with similar variability, and was included primarily as a nonlinear baseline; its default-parameter performance suggests that additional tuning would yield only marginal improvement.

**TABLE 3.** Model comparison using nested five-fold cross-validation (mean AUC ± SD across outer folds).

| Method | AUC (mean) | AUC (SD) |
| --- | --- | --- |
| Ridge | 0.999 | 0.002 |
| SVM (linear) | 0.998 | 0.002 |
| LASSO | 0.989 | 0.015 |
| Random forest | 0.987 | 0.014 |

**FIGURE 6.**
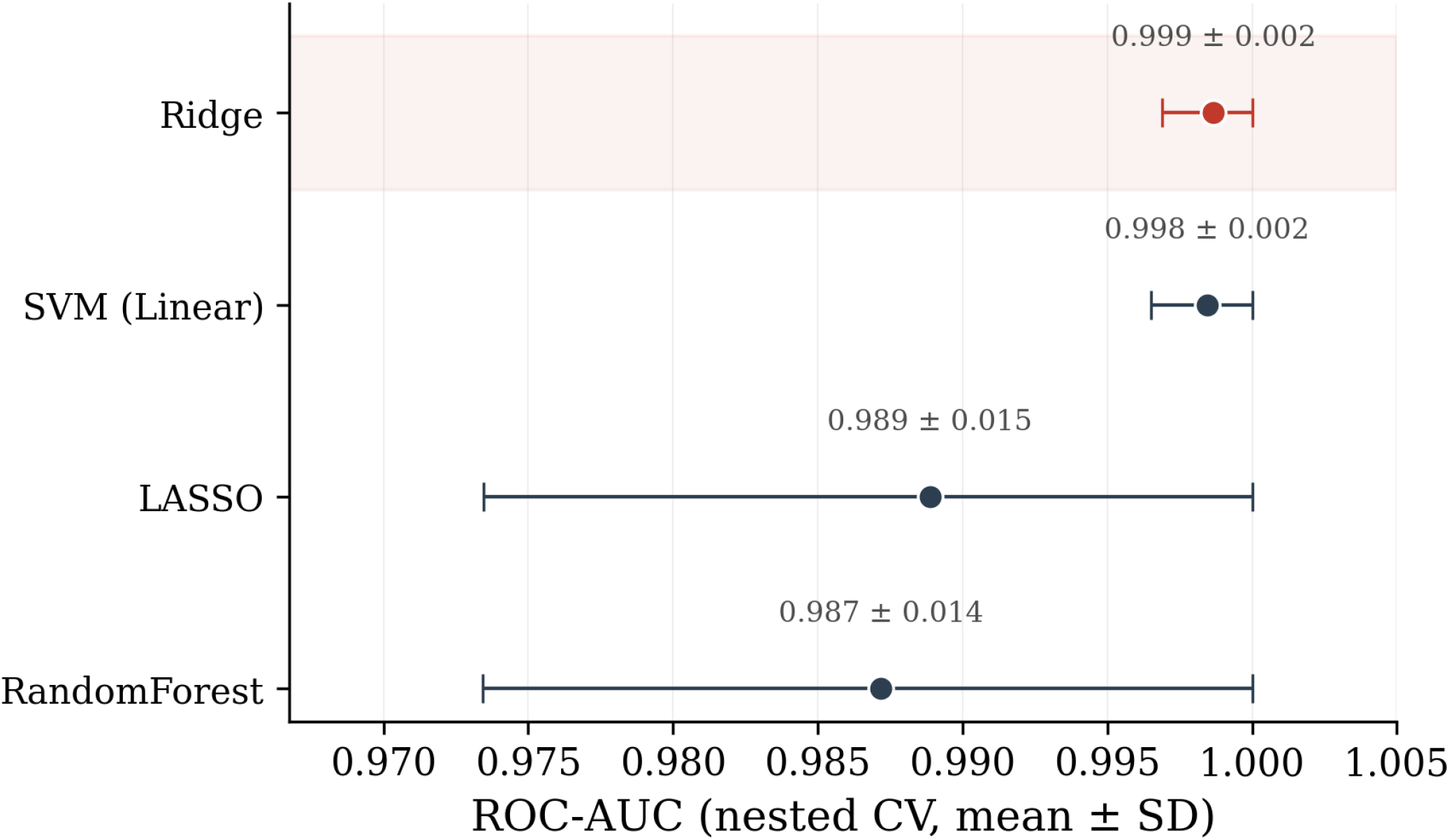
Comparison of four classification methods evaluated using nested five-fold cross-validation. Points represent the mean ROC-AUC across outer folds; horizontal bars indicate ±1 standard deviation.

Overall, a compact set of 29 blood-based features, identified through LASSO within a leakage-free nested cross-validation framework, discriminated lung cancer cases from healthy controls with high accuracy (pooled OOF AUC = 0.990, accuracy = 97.4%, sensitivity = 94.3%, specificity = 99.4%). Permutation testing confirmed that this exceeds chance (*p* = 0.0099), and the six-probe stable core provides further evidence of robustness.

## 4. Discussion

This study presents a methodologically rigorous framework for blood-based lung cancer classification, combining *ℓ*_1_-penalised logistic regression with leakage-free nested crossvalidation, permutation-based inference, and feature stability assessment. Applied to 303 peripheral whole-blood transcriptomic profiles from GSE252168, the framework identified a sparse 29-probe signature achieving a pooled OOF AUC of 0.990, with significance confirmed by permutation testing (*p* = 0.0099).

A central methodological element is the explicit separation of hyperparameter tuning and performance evaluation through nested cross-validation (Varma and Simon, 2006; Cawley and Talbot, 2010). The nested outer-fold AUC of 0.989 ± 0.015 is a conservative and unbiased estimate of within-cohort generalisation performance. Systematic comparison across four classifiers revealed uniformly high performance, indicating a strong, algorithm-independent signal in GSE252168. LASSO achieved comparable discrimination while reducing the feature space to 29 probes (Hastie et al., 2009), and its greater fold-to-fold variability is consistent with the known sensitivity of *ℓ*_1_ estimators to correlated predictors (Haury et al., 2011).

Feature stability analysis highlights an important practical consideration: even near-ceiling performance is accompanied by substantial variability in selected features across resampling splits (Haury et al., 2011; Goksuluk et al., 2016). The six-probe stable core spans functionally diverse categories. *CDC42* is aberrantly expressed in lung cancer and associated with adverse survival (Chen et al., 2012; Zheng et al., 2017), *U2AF1* mutations are among the most frequent splicing factor alterations in lung adenocarcinoma (Ilagan et al., 2015), *RPS15A* is overexpressed in lung adenocarcinoma tissue and promotes tumour cell proliferation (Zhang et al., 2016), and *TTN* carries one of the highest somatic mutation frequencies across solid tumours (Imielinski et al., 2012). Pairwise correlation analysis revealed two dominant co-expression modules (mean |*ρ*| = 0.498), directly illustrating the LASSO tendency to select representatives from correlated groups.

External validation on independent cohorts was not feasible within the scope of this study. A systematic search of the GEO and ArrayExpress repositories identified no additional GPL10558 peripheral whole-blood dataset with a confirmed lung cancer versus healthy control design comparable to GSE252168. The two closest datasets, GSE42834 (Bloom et al., 2013) and GSE135304 (Kossenkov et al., 2019), differ substantially in clinical design and control composition. Peripheral blood gene expression profiles are also highly sensitive to pre-analytical factors such as RNA collection tube type and site-specific laboratory conditions (Asare et al., 2008; Menke et al., 2012), which can introduce systematic expression shifts that compromise cross-cohort comparability. To mitigate the absence of external validation, model development was conducted using a leakage-free nested crossvalidation framework, and permutation testing confirmed that performance significantly exceeded chance.

Several limitations should be acknowledged. First, the 29-probe signature is internally validated within a single dataset, and its clinical generalisability remains unestablished. Second, and most importantly, the study is affected by a structural confound: all cancer cases originated from a single clinical cohort, whereas all healthy controls were drawn from separate population-based biobanks profiled with different RNA collection systems (Tempus and PAXgene tubes). Disease status is therefore partially confounded with cohort of origin and pre-analytical processing, and the possibility that cohort-specific technical variation contributes to the near-perfect discrimination cannot be fully excluded without an independent, matched external cohort; the reported performance should be interpreted with this caveat in mind. A companion study of the same cohort investigates this confound directly using ComBat batch correction and cohort-stratified analysis. Third, clinical covariates (sex, age, smoking status, tumour stage) were not incorporated, as the primary objective was methodological evaluation. Future work will integrate such covariates, examine performance across histological subtypes, and pursue prospective multi-centre external validation using standardised blood collection protocols.

## 5. Conclusion

This study presents a supervised classification framework for high-dimensional blood transcriptomic data that addresses the information leakage and optimistic bias prevalent in the gene expression classification literature. Confining all data-dependent preprocessing and model selection to training partitions enables unbiased performance estimation in settings where the number of features greatly exceeds the number of samples. The LASSO model identified a 29-probe blood-based signature achieving a pooled OOF AUC of 0.990 (accuracy 97.4%, sensitivity 94.3%, specificity 99.4%), with a conservative nested estimate of 0.989 ± 0.015 and permutation-confirmed significance (*p* = 0.0099). A stable six-probe core, including *CDC42, U2AF1*, and *RPS15A*, was selected in all outer folds. The modular framework is broadly transferable to other high-dimensional classification problems across genomics and adjacent domains. Because disease status is confounded with cohort in the present design and no comparable independent GPL10558 whole-blood dataset was identified for external validation, prospective multi-centre validation under standardised pre-analytical conditions remains the critical prerequisite for clinical translation.

## Data Availability

All data analysed in this study are publicly available online from the Gene Expression Omnibus (GEO) under accession number GSE252168 (https://www.ncbi.nlm.nih.gov/geo/query/acc.cgi?acc=GSE252168). No new data were generated.

## Data availability

The gene expression dataset analysed in this study is publicly available from the Gene Expression Omnibus under accession number GSE252168.

## Ethical Statement

This study analysed publicly available, fully de-identified human gene expression data obtained from the Gene Expression Omnibus (accession GSE252168). No new data were collected from human participants, and all analysed data were anonymised and publicly accessible; therefore, no additional institutional review board approval was required for this secondary analysis.

